# Understanding the effect of pulse width on activation depth in TENS: A computational study

**DOI:** 10.1101/2024.04.10.24305618

**Authors:** Alexander Guillen, Dennis Q Truong, Yusuf O. Cakmak, Sheng Li, Abhishek Datta

**Author notes:** Authors 1 and 2 have equally contributed to the article (Alexander Guillen, Dennis Q Truong). (AD).

## Abstract

**Background:** Transcutaneous electrical nerve stimulation (TENS) has been a commonly used modality to relieve aches and pain for over 40 years. Commercially available devices provide multiple therapy modes involving a different combination of frequency and pulse width with intensity. While frequency sets sensation, intensity helps determine tolerability, longer pulse width is reported to induce a feeling of deeper stimulation. In fact, longer pulse width has been empirically shown to deliver current into deeper tissues, but in context of other electrical stimulation modalities. The goal of this study was to unpack the relationship between pulse width and activation depth in TENS.

**Methods:** A highly realistic, anatomically-based, 3D finite element model of the forearm was used to simulate the electric field (E-field) distribution, as the pulse width is varied. A typical titration-guided mechanism was used to obtain the strength-duration (S-D) curves of a sensory McIntyre-Richardson-Grill (MRG) axonal model simulating the pain-transmitting A-delta fibers. The pulse widths tested ranged from 30 μs to 495 μs.

**Results:** As expected, shorter pulse widths required more current to achieve activation, resulting in a larger E-field. The S-D curve of the target median nerve indicates a rheobase of 1.75 mA and a chronaxie of 232 µsec. When the applied currents are the same, shorter pulse widths result in a smaller volume of tissue activated (VTA) compared to the longer pulse widths. A 21 fold difference in VTA was found between the longest and shortest pulse widths considered. We observed a linear relationship between pulse width and activation depth for the conditions tested in the study.

**Conclusion:** Our findings highlight the impact of pulse width on activation depth. While choice of a given therapy mode is usually based on an *ad-hoc* desirable sensation basis, medical professionals may consider advocating a certain therapy mode based on the depth of the intended target nerve.

## Introduction

Pain is the body’s response to injury or illness. Usually, the body heals and the pain goes away, but for many people, the pain persists long after the cause has gone away. The Centers for Disease Control and Prevention (CDC) estimates that 20.4% (50 million) of American adults suffer from chronic pain and, of which 8% (19.6 million) live with high-impact chronic pain [1]. Chronic pain is defined by the International Association for the Study of Pain (IASP) to be pain persisting beyond typical tissue healing time, which is generally considered to be 3 months [2]. Common types of chronic pain include back, headache, joint, neck, hip, and osteoarthritis pain [3]. Although treatment typically includes pharmacological approaches, one non-pharmacological and non-invasive option recommended by some clinicians for its convenience and effectiveness is Transcutaneous electrical nerve stimulation (TENS) therapy [4].

Studies suggest that TENS helps reduce pain via peripheral and central mechanisms. In the central system, TENS activates the sites in the spinal cord and brainstem that use opioid, serotonin, and muscarinic receptors; and through peripheral mechanisms — opioids and ɑ-2-noradrenergic receptors that intervene in the induction of analgesia [5]. Using small battery-powered devices, the modality typically delivers biphasic, symmetric or asymmetric, rectangular or square pulses through cutaneous electrodes positioned near the painful area [6]. They can be applied with varying frequencies, from low (< 10 Hz) to high (> 50 Hz), or mixed frequencies [7]. In general, higher-frequency stimulation is delivered at sensory intensity, and low-frequency stimulation is delivered at motor intensity [6]. At sensory intensity, patients may experience strong but comfortable sensations without movement contractions, whereas at high intensity they can feel painless motor contraction.

The early evolution of TENS has been characterized by a faster rate of development of clinical applications rather than determining optimal parameters [8]. This has been compounded by the fact that use for low back pain was “grandfathered” in the United States. As TENS for low back pain was marketed prior to the 1976 medical device regulation act, it was allowed to stay in commerce, and therefore TENS efficacy for low back pain was never “premarket approved”. Newer devices could thereby, obtain marketing “clearance” based on demonstrating equivalence to prior devices based on technology (stimulation parameter) comparison. As there was no motivation for device manufacturers to develop proper clinical utility and generate high quality efficacy data, clinical evidence has continued to be debated [3,4]. Newer indications such as TENS for migraine and sinus pain have however demonstrated definitive clinical utility [9–11]. TENS devices are considered medium risk (Class 2) devices and are available for both prescription and OTC use.

Commercially available TENS devices for peripheral pain provide multiple therapy modes with each mode delivering a different combination of frequency and pulse width with intensity. Users are asked to screen through available modes and settle on the mode that provides the “most desirable sensation / comfort”. While frequency selection allows to set desired sensory or motor contraction, intensity generally maps to tolerability, longer pulse width is suggested to induce a feeling of deeper stimulation.

Studies exploring the effects of pulse width / duration over the years, have mostly studied physiological responses and not the exact relationship to activation depth in TENS. Li and Bak 1976 [12] showed that isolated excitation of different nerve groups (motor, sensory, pain-conducting fibers) may be easier with a short duration pulse. Effects on pulse width on the arm have reproduced basic relationships between pulse duration and current intensity found in prior literature [13, 14]. Specifically, Alon et al., 1983 [8] demonstrated that pain thresholds mediated by pain-conducting fibers in healthy subjects resulted in reducing pain thresholds (350 mA - 30 mA) as pulse duration was increased (5 μs - 1000 μs). Further, stimulus pulse width may also be used to selectively recruit fibers of different sizes [15]. Some efforts do however come close in context of related electrical stimulation modalities. Increasing pulse width was empirically shown to improve current penetration by reaching distant muscles from surface electrodes in neuromuscular electrical stimulation (NMES) [16]. In the context of invasive DBS, increasing pulse width has been shown to lead to activation at greater distances from electrode center (or deeper stimulation) [17, 18]. Further, long pulse width stimulation has been shown to penetrate and activate deeper muscles in functional electrical stimulation [19, 20].

The goal of this computational study was to investigate the effect of the pulse width in TENS on the arm. A high anatomically realistic finite element model was used to simulate the induced electric field (E-field) distribution. The E-field is then coupled to a sensory neuron model given TENS’s efficacy is predicated on providing pain relief by exciting sensory nerves. We evaluated strength-duration (S-D) curve and volume of tissue activated (VTA). The VTA map was related to pulse width to provide insight on the effect of pulse width on activation depth.

## Methods

### Geometry setup

The computational study was performed using Sim4Life (V7.0.1, Zurich MedTech, Zurich, Switzerland) incorporating NEURON solver (v7.2.3.12730). Sim4Life is a simulation platform that combines human phantoms with relevant physics solvers for analyzing real-world biological problems. The model geometry used in the study corresponds to the right arm of the Yoon-sun V4-0 dataset [21]. The model incorporates high resolution data (0.1 x 0.1 x 0.2 mm) making it possible to resolve and thereby segment nerves, arteries, veins, and other small structures. Importantly for this study, it includes all major nerve trajectories from the cranium and spinal cord to internal organs and muscles and has been used in other peripheral nerve stimulation studies [22, 23]. The relevant tissue properties for this study are presented in **Table 1** and based on the IT’IS material parameter database [24]. Precisely, the model in **Figure 1A** consists of a pair of stimulation electrodes placed on the wrist with the goal of targeting the medial nerve. The electrodes are placed along the length of the medial nerve and only serves as an exemplary placement to study the effect of pulse width. The major underlying layers that comprise the model are further indicated in **Figures 1B** and **1C**. The electrode and interfacing gel combination have a radius of 3 mm with a combined total thickness of 2 mm.

**Figure 1:**
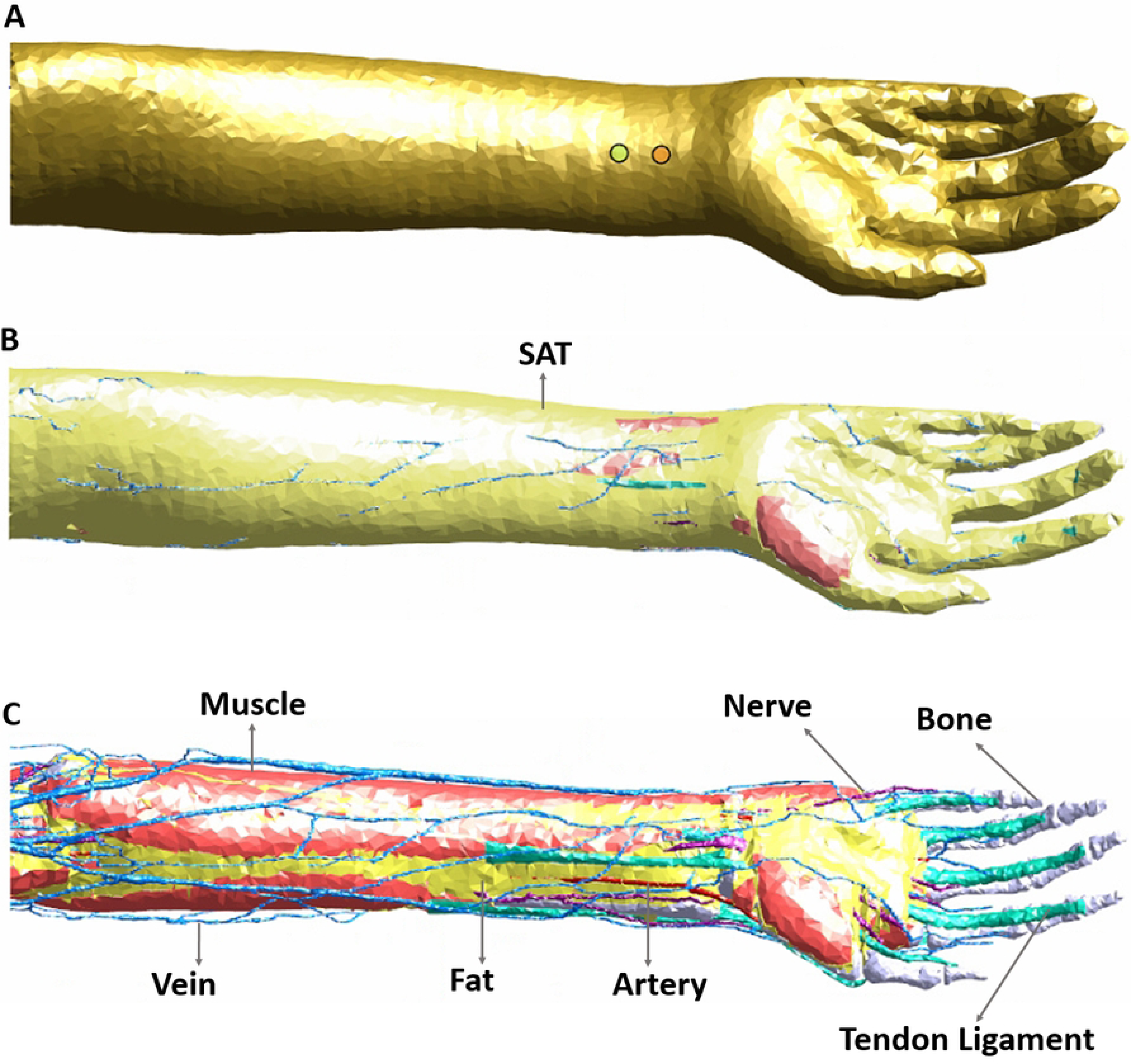
Arm geometry and tissue composition. **(A)** indicates the position of the stimulation electrodes targeting the median nerve. **(B)** indicates the subcutaneous adipose tissue (SAT) layer. **(C)** indicates other underlying tissues such as muscle, nerves, bone, etc. Note: Refer to **Table 1** for all tissues considered in the model and their corresponding electrical conductivities.

**Table 1:** Tissue electrical conductivities (S/m) used in the model.

| Tissue | Conductivity | Tissue | Conductivity |
| --- | --- | --- | --- |
| Skin | 0.1482 | Tendon/ligament | 0.3675 |
| Bone (Cancellous) | 0.0804 | Muscle | 0.4610 |
| Bone (Marrow) | 0.1797 | Air | 0 |
| Bone (Cortical) | 0.0063 | Blood | 0.6624 |
| Fat | 0.0776 | Nerve | 0.3475 |
| Subcutaneous adipose tissue (SAT) | 0.0776 | Gel electrode | 1.7 |

### Nerve trajectories

The intended stimulation region and corresponding nerve anatomy is shown in **Figure 2A**. While the regions of interest are areas in immediate proximity to the stimulation electrodes, overall visualization of anatomical details in the considered geometry is helpful, to relate to induced E-field and VTA plots. As is known, five specific nerves appear from the cords as the terminal branches of the brachial plexus: musculocutaneous, axillary, radial, median and ulnar nerves. The musculocutaneous nerve provides motor innervation to the muscles of the anterior compartment of the arm [25]. The median nerve (comprising C6-T1 spinal roots) predominantly provides motor innervation to the flexor muscles of the forearm and hand [26]. The radial nerve innervates most of the skin of the posterior forearm, the lateral dorsum of the hand, and the dorsal surface of the lateral three and a half digits. Lastly, the ulnar nerve carries both sensory and motor fibers and supplies sensory cutaneous innervation to the medial forearm, medial wrist, and medial one and one-half digits [25].

**Figure 2.**
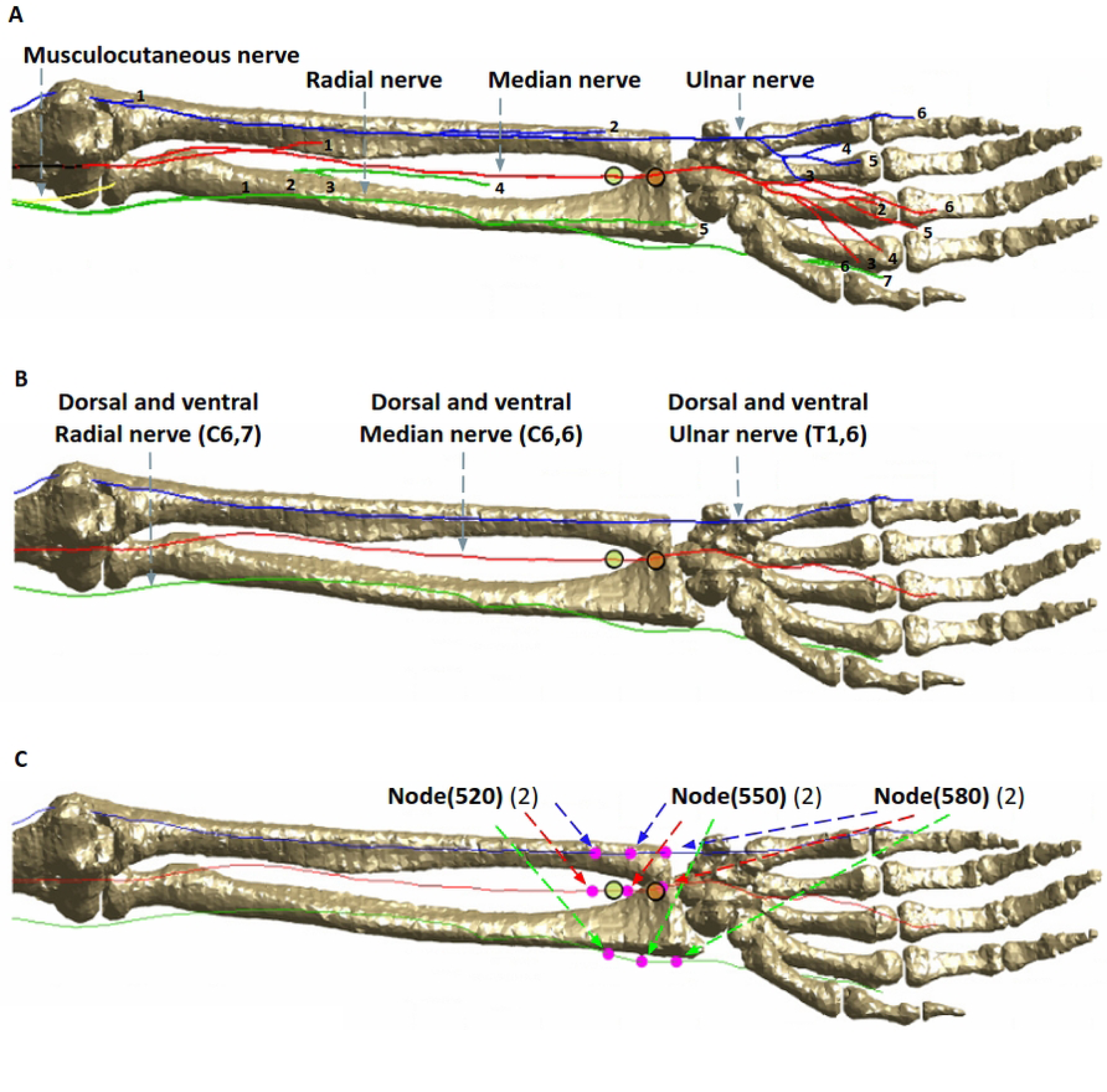
Nerve anatomy in the arm and locations evaluated. **(A)** Available terminal branches of the brachial plexus nerves (musculocutaneous, radial, median, and ulnar) highlighting anatomical detail in geometry. **(B)** The longest nerve trajectories were subsequently considered for simulation. For instance, (C6,6) refers to the nerve segment in the cervical section (C6) with 6 being the specific trajectory number. The ulnar nerve has 1,2,3,5,6 trajectories in the thoracic section but we consider the longest trajectory (6), or (T1,6). **(C)** Point sensor locations. The nodes (520,550,580) indicate the exact location of simulation data collection.

The longest trajectories of the ventral and dorsal roots of C6 and T1 were considered for analysis here (**Figure 2B**) due to the expected direct influence on the mid-forearm - based on electrode locations. The nerve depth from skin in contact with electrodes to the median nerve is about 5.5 mm, the radial nerve is about 13.1 mm, and the ulnar nerve is 14.9 mm. The three “point sensor” locations along the nerve trajectory used for collecting the simulation data (i.e. nodes 520, 550, and 580) are illustrated in **Figure 2C**. There were therefore 18 point sensors considered: 9 (ventral rami) and 9 (dorsal rami).

### Injected current

The low-frequency electromagnetic (EM LF) - Ohmic Quasi-Static module, a rectilinear LF solver, was used to simulate TENS on the arm at multiple pulse widths (30, 88, 146, 262, and 495 μs) corresponding to the range typically available in commercial TENS stimulators. Dirichlet boundary conditions were applied as 2.38 V and - 2.38 V at the anode and cathode corresponding to 5 mA of current flux calculated on the electrodes. Other external boundaries were electrically insulated (i.e. normal current density = 0). The Ohmic quasi-static field

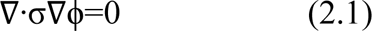

was solved with the aforementioned boundary conditions for the electric potential distribution [27–29]. The injected current was then re-calculated based on the titration factor (see section on Titration mechanism) to visualize differences in the E-field between the different pulse widths.

### Neuron model and additional simulation considerations

TENS is known to stimulate sensory nerves, suppressing the pain signals being sent to the brain to give user relief. We therefore considered the sensory McIntyre-Richardson-Grill (MRG) neuron model [30,31], with a diameter of 5 μm, to simulate the effects of pain-transmitting nerve fibers. The MRG model is based on a double-cable representation of the axon that allows separating electrical representations of the myelin and underlying internodal axolemma. The model has been used for neural predictions in a variety of applications [18, 32, 33]. For simulating TENS using the titration mechanism, the modulation pulse type was set to bipolar with a unitless amplitude of 1 and an interphase interval of 0.1 ms while varying the pulse widths. The duration and time step were set to 3.5 ms and 0.0025 ms respectively. Running a simulation for the aforementioned nodes of interest at one pulse width took approximately 4 hours using 64 threads on a workstation with the following specifications: AMD Ryzen Threadripper 3970X 32-Core Processor, 3.70 GHz CPU speed, and 192 GB installed RAM.

### Titration mechanism

Titration involves stimulating an axon with a series of pulses of *increasing* intensity to find the threshold at which a single action potential is generated in excitable cells. This method introduces an additional scaling factor that is titrated until a response can be detected within the stimulated region [34, 35]. Thus, the excitability threshold (I_T_) is the product of the current applied to the cellular membrane of the axon, the aforementioned titration factor (T), and the modulating pulse (a(t)):

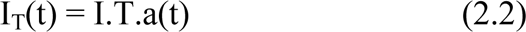

The T parameter is considered as a scaling factor to indicate proportion or a multiple of the actual modulated current needed to generate an action potential. Potential and current can be used interchangeably here for measuring the stimulus strength.

### Volume of Tissue Activated

The volume of tissue activated (VTA) was used to compare stimulation differences caused by changes in pulse width. The VTA around each electrode contact relied on the concept of activation function (AF), which was calculated from the eigenvalues of the Hessian matrix [36]. Each eigenvalue of the Hessian matrix represents the second partial derivative of the electric potential along the respective eigenvector. A multi-step process was used to determine the VTA due to pulse width variation. This involved determining the excitability threshold (I_T_), using the corresponding electric potential to calculate the AF, and subsequently utilizing the AF to determine the VTA.

## Results

### Electric Field (E-field)

The induced *surface* E-field plots due to the shortest and the longest pulse widths, calculated at their corresponding titration factors (see **Table 2**) and plotted to the same scale, are included in **Figure 3**. The plots reveal that the smaller pulse width (30 μs) induces a larger E-field (max: 456 V/m) — covering a larger area of the arm. On the other hand, the induced E-field due to 495 μs is less diffuse, more focused, and has lower magnitude from the same pair of electrodes. However, this is intuitively expected, as the plots are generated at their respective stimulation threshold, so the 30 μs E-field is the result of 19.7 mA and the 495 μs E-field is the result of injecting 1.75 mA. The overall spatial profile resembles a stretched ellipse with the major axis along the line connecting the stimulation electrodes. The ulnar and radial nerves that are farther away from the electrode sites receive less E-field (∼ 0-132 V/m). With longer pulse width, the induced E-field profile in the immediate vicinity of the electrode sites indicates a restricted hot-spot with dramatic fall-off (∼40-280 V/m) and approximately 0-23 V/m for the rest of the nerves.

**Figure 3.**
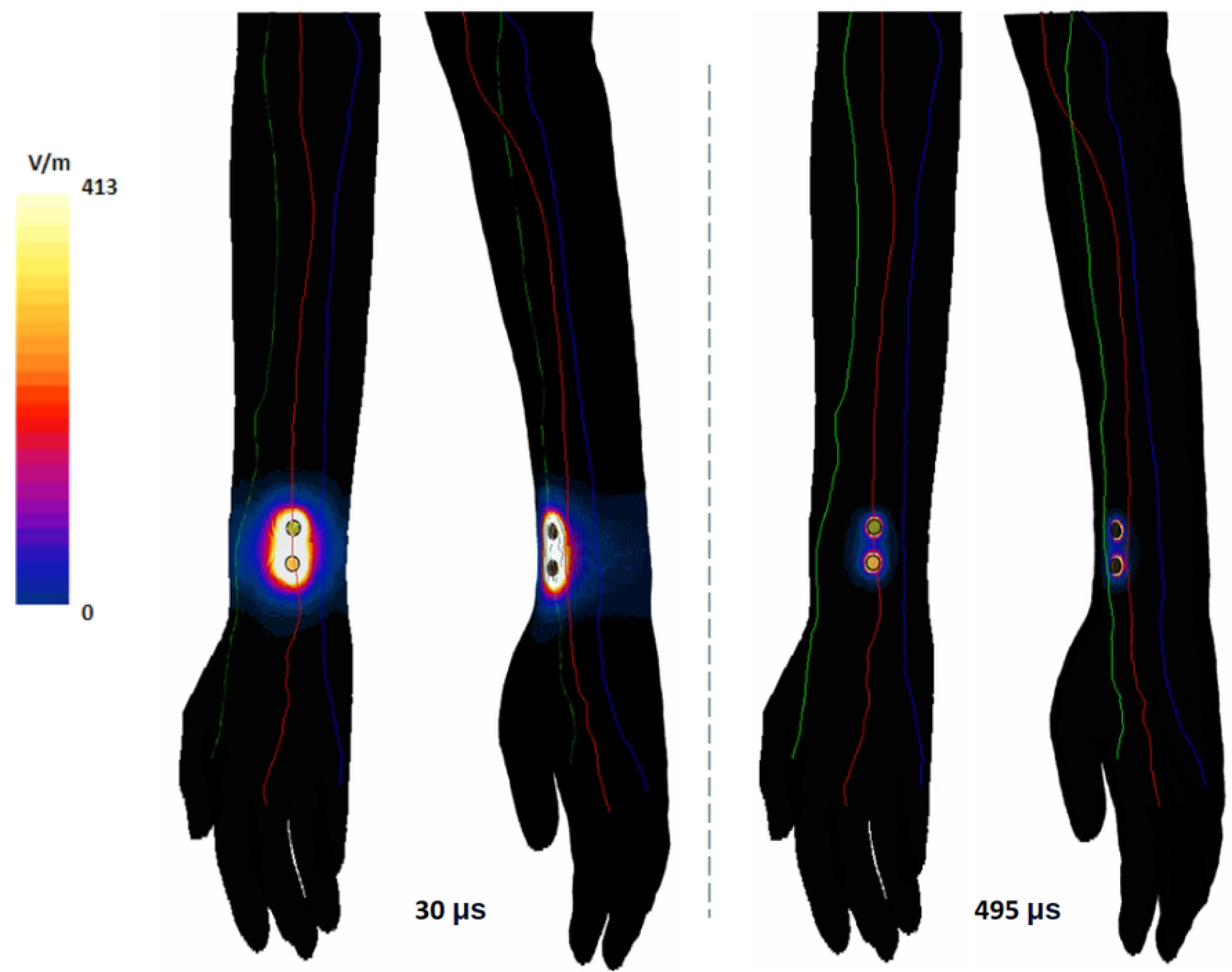
Surface plot of induced E-field on the arm due to the shortest and longest pulse widths considered. The E-field was calculated based on the current due to their corresponding titration factor: for 30μs, input current was 19.7 mA and for 495μs, 1.75mA.

**Table 2.**
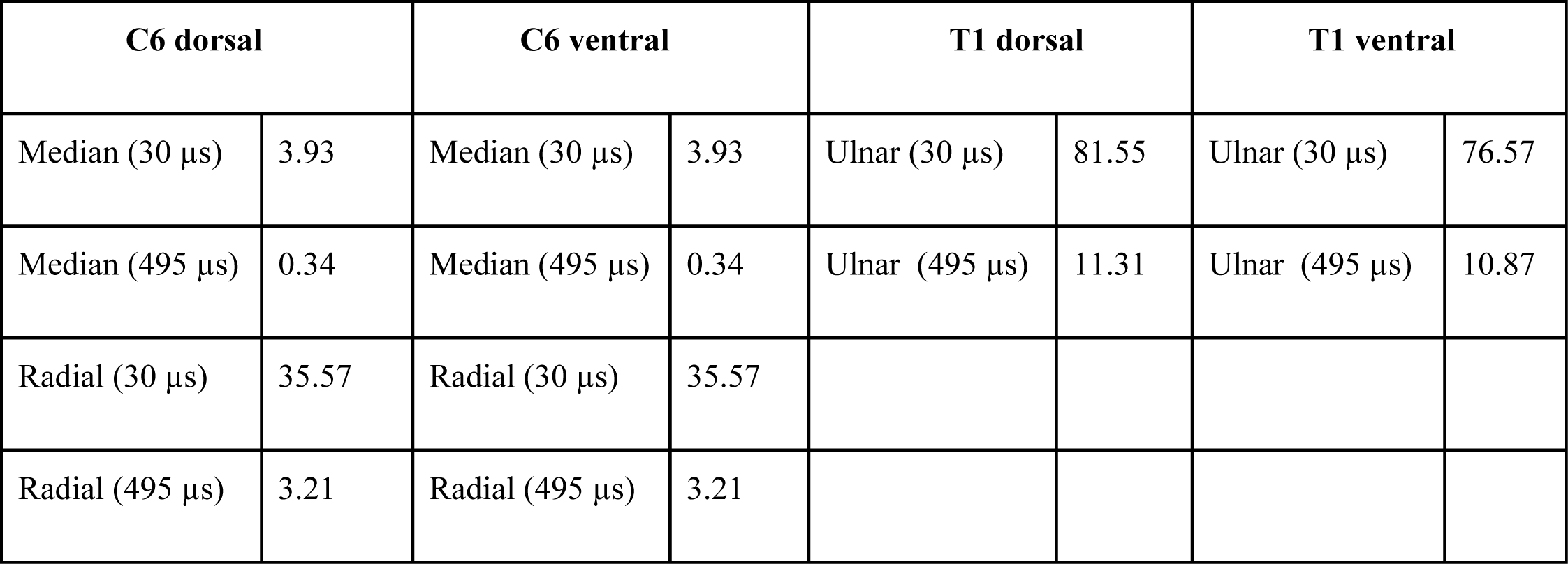
Titration factor at chosen sensor locations. Titration factor for the shortest (30μs) and longest (495μs) pulse duration at sensor nodes (520,550,580). As expected, minimum titration factors were needed for the superficial median nerve with the highest values needed for the ulnar nerve.

### Titration Factor

As previously stated, the titration technique was employed to find the threshold potential of membrane depolarization. **Table 2** notes the individual titration factors needed for the roots (C6 and T1). As expected, the titration factor is substantially smaller for the 495 μs pulse in comparison to the 30 μs pulse. While the titration factors of dorsal and ventral sections for the C6 roots are the same, they differ somewhat for the T1 roots. Further, as anticipated, minimum titration factors were needed in the branches of the median nerve due to the proximity to the electrode sites.

### 3.3 Strength-Duration Curve

The resulting strength-duration (S-D) curve of the median nerve under electrical stimulation is shown in **Figure 4**. For the range of 30-495 μs considered here, the rheobase was found to be ∼1.75 mA with a chronaxie of ∼232 μs. Consistent with the theory, the curve tends to flatten out with longer stimulus duration (or pulse width).

**Figure 4.**
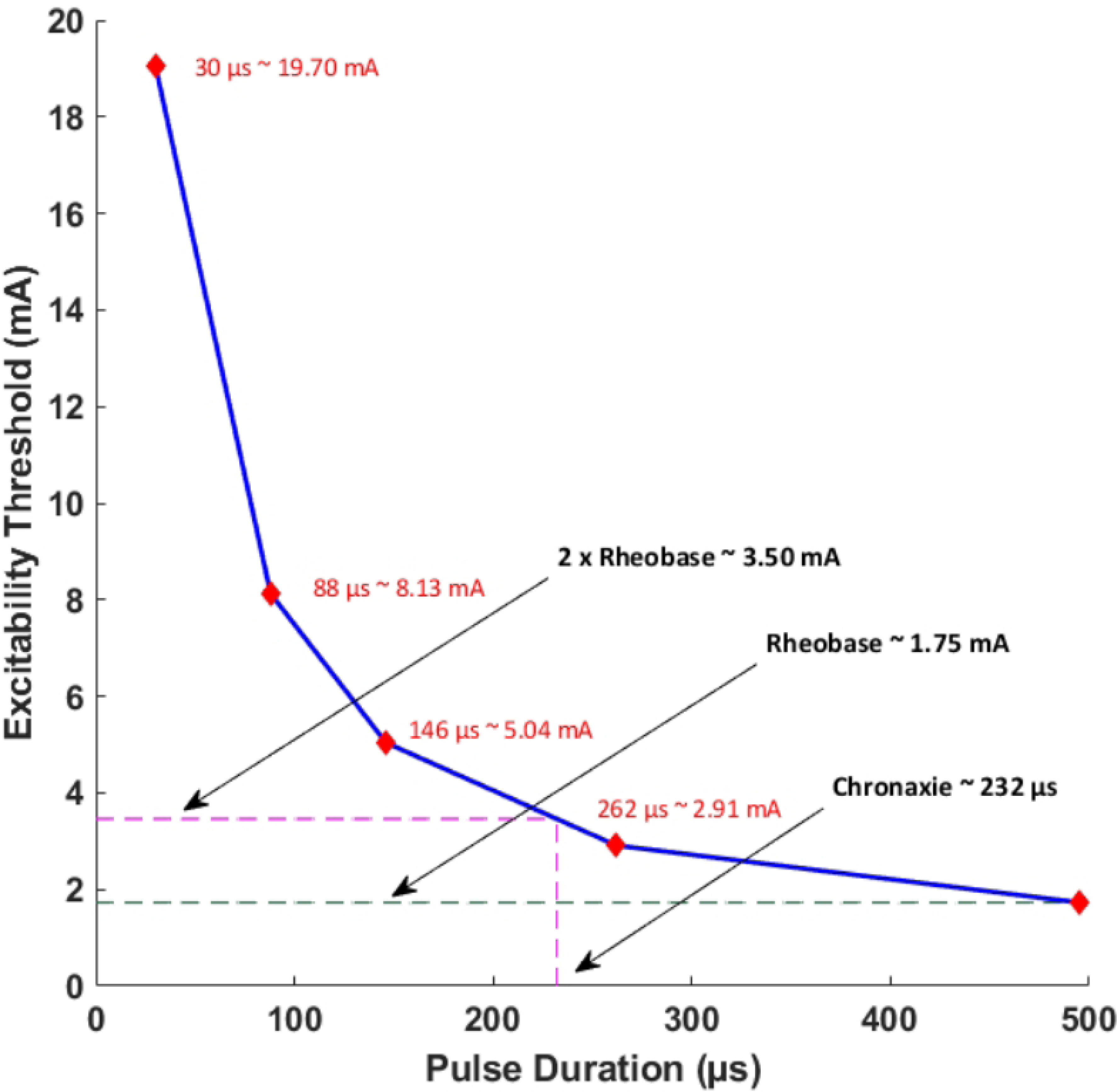
Strength-Duration (S-D) curve of the target median nerve. A pulse width range of 30-495 μs was considered in the study. The corresponding excitability threshold for each pulse width is noted along the curve.

### 3.4 Volume of Tissue Activated (VTA)

The VTA maps illustrate isosurface plots derived from the absolute value of Hessian matrix eigenvalues of the electric potential. Since the Hessian matrix is essentially a matrix of the second partial derivative of the electric potential, it enables determination of the classic activating function in 3D [37, 38]. To facilitate a direct comparison across the range of pulse widths considered, we plotted the VTA maps at one common current value-i.e. the average threshold current (10.73 mA) spanning the shortest and longest pulse widths (**Figure 5**). As expected, the shortest pulse width requires the highest threshold to activate tissue near the input source and is approximately a factor of 11 higher with respect to the longest pulse width (8.8 e^5^ / 7.82 e^4^). The estimated VTA for the shortest and widest pulses were 118.72 mm^3^ and 2,586.24 mm^3^ respectively, indicating a VTA ratio of 21.2.

**Figure 5.**
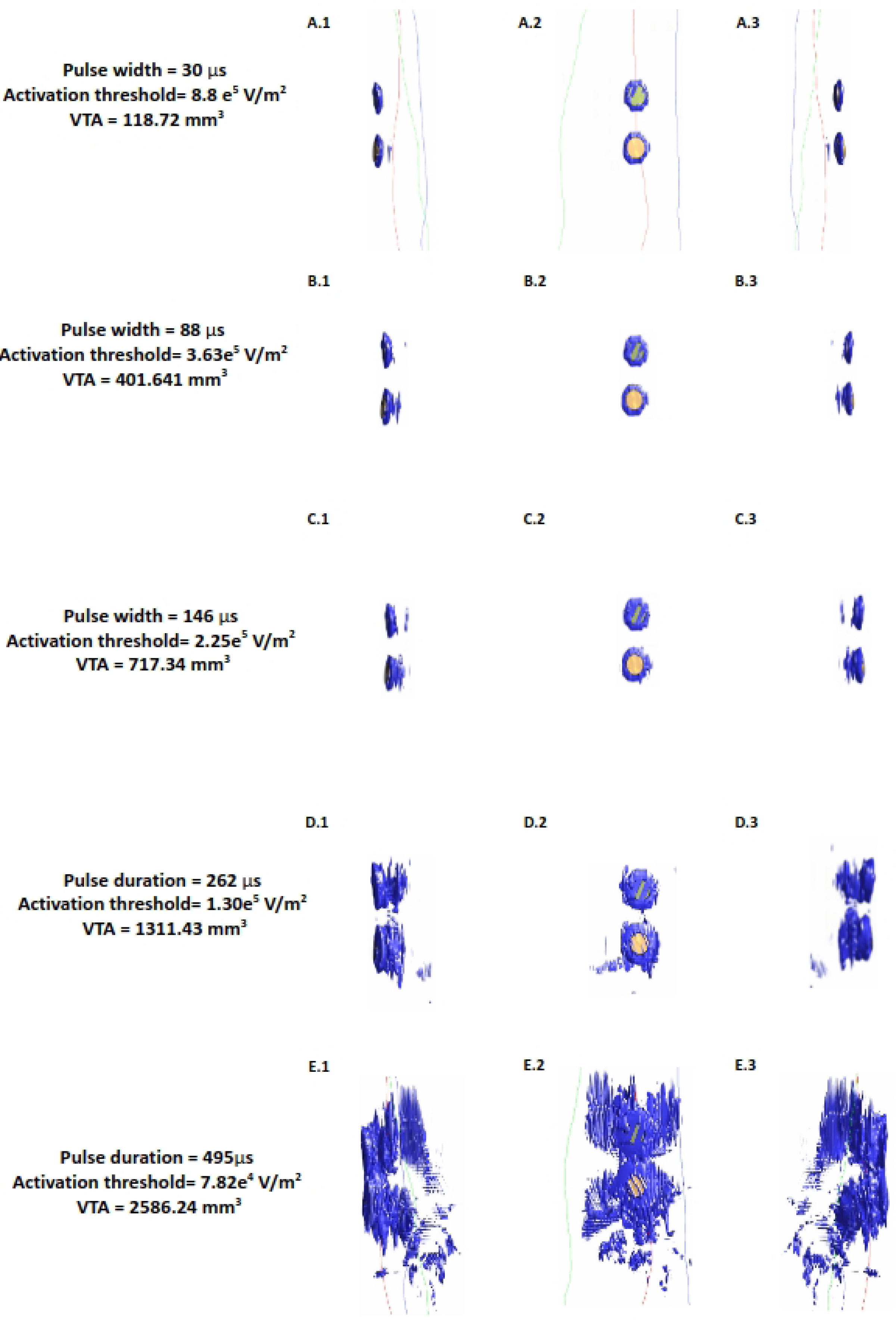
Median nerve activating function (AF) and volume of tissue activated (VTA) at a fixed current amplitude (10.73 mA) Figures show the isosurface plots of the second spatial derivative of electric potential at the AF threshold for the median nerve. Each row is the result of a simulated pulse width in ascending order (30, 88, 146, 262, 495 μs).

The value of this study is exemplified by observing the shape and pattern of the VTA maps. While VTA investigation in invasive applications such as DBS reveal uniform “blobs’ ’ around the electrode contacts reflecting *one* brain region [17, 39], the maps here are scattered and irregular, due to varying complex anatomy. This is only captured due to the realistic arm geometry considered here. The VTA maps also help visualize the influence of pulse width on activation depth. The plots in the first column indicate that for pulse widths up to 146 μs, it is not possible to recruit the deeper radial and the ulnar nerves. At the longest pulse width, there is some activation at the levels of the deeper nerves of the arm.

### Influence of the pulse width with respect to activation depth

To understand the influence of pulse width on activation depth, we plotted predicted VTA with respect to the varying pulse widths considered (**Figure 6**). We note a linear relationship for the range of pulse widths considered here.

**Figure 6.**
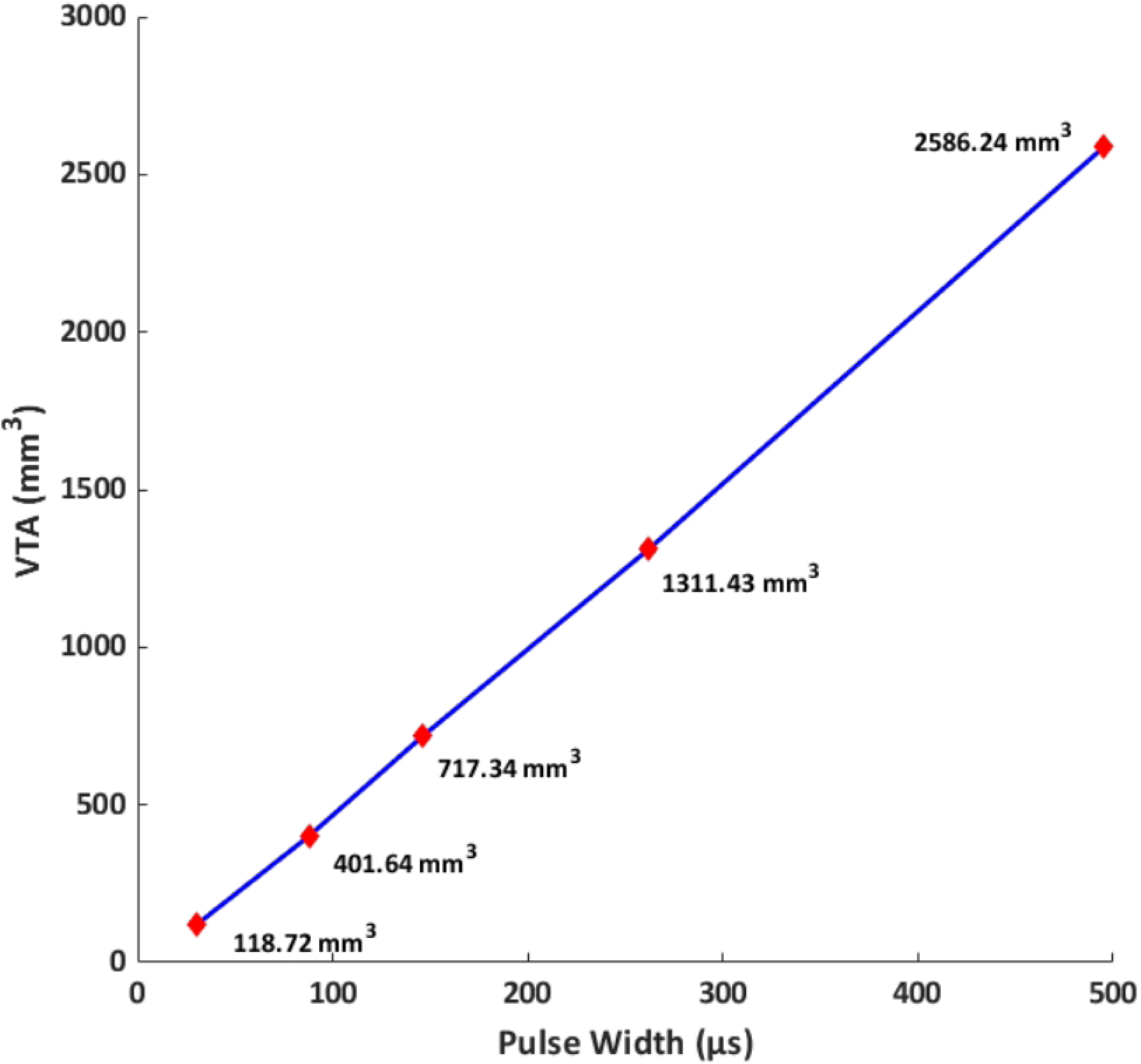
Volume of tissue activated (VTA) vs pulse width at a fixed current amplitude (10.73 mA) VTA is used as a surrogate for activation depth. For the range of pulse widths and conditions considered here, the relationship is linear.

## Discussion

The central aim of this study was to unpack the relationship between pulse width and activation depth during TENS on the arm. Prior electrical stimulation studies using other modalities have empirically shown that wider pulses can recruit deeper targets. Using a highly detailed 3D model, we provide enhanced visualization of available geometry, induced E-field profile, VTA maps, and relationship to activation depth, for the first time.

The S-D curves for nerve stimulation have clearly established its shape over numerous investigations dating back to the 30’s. Given the dominant electrical capacitance of the neural membrane, S-D curves expectedly follow a capacitor discharge curve. A TENS practitioner can therefore readily use the inverse relationship between intensity and pulse width to make an informed stimulation strategy choice. Now longer pulse widths at the same current would lead to more charge delivered across the membrane- presumably translating to deeper stimulation. However, the exact relationship has not been explored previously in TENS. Further for non-invasive electrical nerve stimulation applications, lowering pulse width in order to deliver higher current intensity is limited to the intensity at which the user can comfortably receive stimulation [40]. This restriction is however not applicable for invasive delivery [18] as stimulation does not have to navigate superficial cutaneous sensation.

The linear relationship between VTA and pulse width observed in our simulations indicates that longer widths would lead to deeper activation. We note that our observations are restricted to only the pulse widths and the concomitant geometry (arm) considered here. We expected the relationship to asymptote at higher pulse widths as the excitability threshold reaches rheobase. While the excitability threshold in the S-D curve (**Figure 4**) follows a hyperbolic or exponential decay similar to classical equations (Weiss-Lapique and Lapique-Blair [41, 42]), the VTA expands with a similar convexity resulting in a *net* linear VTA - pulse width relationship. We suspect this is due to (1) volume being cubic and (2) the Hessian of voltage dropping exponentially away from the electrodes. There are several limiting assumptions to the plot. We are considering only the magnitude of the Hessian, which does not account for orientation / alignment with any possible axon. The AF thresholds were calibrated for the median nerve A-delta fiber running along the length of the arm; other nerve orientations and fiber types would be expected to respond differently. Further, heterogeneous tissues cause spikes in E-field and AF at material boundaries.

TENS efficacy is likely predicated upon a net effect of stimulating multiple underlying nerves of various types and not just the A-delta fiber considered here. We also note that we simply used the median nerve in this study as a test nerve to explore relationships between E-field, strength, duration, activation depth, etc. However, the choice of A-delta fiber is rational as nociceptors generally transmit noxious stimuli through A-delta and C-fiber nerves. Further, it is known that the C-fiber afferents carry slow sensations associated with aches, whereas the A-delta afferents are associated with fast sensations such as sharp pain. We expect our main results to also hold for C-fiber median nerves with differences in rheobase and chronaxie values. Given the type of pain felt and if one were to know underlying nerve depth [43], one could potentially start with a suitable pulse width. In reality however, pre-programmed therapy modes (combination of pre-set frequency and pulse width) are provided in TENS devices, limiting full flexibility to the user in parameter selection. Therefore, the best approach continues to be to try all modes first, and in each case, titrating intensity to the strongest possible but at a level that is comfortable. If deeper pain relief is desired, patients should then pick the next mode with longer pulse width while maintaining the frequency and intensity from the prior mode.

The strength of our modeling process in simulating TENS on the arm is the usage of a highly realistic model. Previous 3-D FEM approaches have either used idealized geometries such as a cylindrical arm [44–46] or derived from 2-D anatomical images by extruding geometry [35] and limiting to certain cross sections [32]. The modeling methodology applied here, from the geometry, applying EM simulations into a dynamic Neuron solver, using the MRG model, and subsequently using titration analysis, mimics the one employed in the context of magnetic stimulation (MS) [22]. The simulation setup used in the aforementioned MS study has been further validated using clinical experiments. Specifically, numerically estimated latencies and waveforms were in agreement with the empirical measurements on subjects undergoing MS on the arm [23]. The only difference to our simulation is the application of electrical stimulation and thereby, related governing equation. However, the governing equation is a standard equation used to predict induced current in volumetric media and has been validated in other applications [47, 48]. Further, the MRG model has been shown to generate accurate predictions for TENS specifically, compared to active cable and mammalian nerve models [27, 35]. Taken together, we expect the main conclusions of this study to be robust.

There are practical limitations to increasing the pulse width at the same current intensity to increase activation depth, namely battery life. The chronaxie is usually considered the most efficient pulse width choice for conserving the pulse generator’s battery life and is naturally a key factor in invasive applications [49]. With modern day TENS devices powered by high capacity small rechargeable batteries, this is not much of a concern. However, as mentioned above, we expect the linear relationship to ultimately change and plateau. Promising solutions such as coupling TENS with a nerve cuff to facilitate activation of deeper nerves has been proposed [35]. However, nerve cuff involves surgery and moreover, such solutions are still being developed and not currently available to the practitioner or the patient.

Computational modeling and simulation such as the one reported here is now increasingly used across a range of stimulation modalities, from optimizing delivery, performing safety analysis, to supporting device design / development [29,39 50-53]. Furthermore, these predictions have helped in elucidating stimulation parameter choices, understanding mechanism of action, explaining stimulation outcome, and thereby advancing stimulation administration in general [11, 54–57]. We expect this study on the arm to guide researchers in performing future explorations on other body parts, determine ideal pulse width range for target nerve of interest, attempt validation using TENS similar to the one performed in MS [23], and investigate new TENS delivery approaches.

## Data Availability

All relevant data are within the manuscript and its Supporting Information files.

## Author Contributions

AD developed the concept idea. AG and DQT performed the modeling and related post-processing. AG, DQT, and AD analyzed the results and prepared the initial manuscript draft with important intellectual input from YC and SL. All authors confirmed the overall methodology, contributed to the final manuscript text, and approved the submitted version.

